# Machine learning-based single cell and integrative analysis reveals that baseline mDC predisposition predicts protective Hepatitis B vaccine response

**DOI:** 10.1101/2021.02.22.21251864

**Authors:** Brian Aevermann, Casey P. Shannon, Mark Novotny, Rym Ben-Othman, Bing Cai, Yun Zhang, Jamie C. Ye, Michael S. Kobor, Nicole Gladish, Amy Lee, Travis M Blimke, Robert E. Hancock, Alba Llibre, Darragh Duffy, Wayne C. Koff, Manish Sadarangani, Scott J. Tebbutt, Tobias R. Kollmann, Richard H. Scheuermann

## Abstract

Vaccination to prevent infectious disease is one of the most successful public health interventions ever developed. And yet, variability in individual vaccine effectiveness suggests a better mechanistic understanding of vaccine-induced immune responses could improve vaccine design and efficacy. We have previously shown that protective antibody levels could be elicited in a subset of recipients with only a single dose of the hepatitis B virus (HBV) vaccine. Why some, but not all, recipients responded in this way was not clear. Using single cell RNA sequencing of sorted innate immune cell subsets, we identified two distinct myeloid dendritic cell subsets (NDRG1-expressing mDC2 and CDKN1C-expressing mDC4), the ratio of which at baseline (pre-vaccination) predicted immune response to a single dose of HBV vaccine. Our results suggest that the participants in our vaccine study were in one of two different dendritic cell dispositional states at baseline – an NDRG2-mDC2 state in which the vaccine elicited an antibody response after a single immunization or a CDKN1C-mDC4 state in which the vaccine required two or three doses for induction of antibody responses. Genes expressed in these mDC subsets were used as an approach for feature selection prior to the construction of a predictive model using supervised canonical correlation machine learning. The resulting model showed an improved ability to predict serum antibody titers in response to vaccination. Taken together, these results suggest that the propensity of circulating dendritic cells toward either activation or suppression, their “dispositional endotype”, could dictate response to vaccination. The fact that these mDCs could be modulated via TLR stimulation could guide progress towards design of effective single dose vaccination strategies.

## Introduction

Vaccination as a general strategy to prevent infectious disease has been one of the most effective public health measures since its conceptualization and implementation by Edward Jenner in the 18^th^ century, and has resulted in the complete eradication of smallpox, the near elimination of polio, and a dramatic reduction in the incidences of measles, mumps and other common diseases. In contrast to these successes, several notable failures in the development of effective vaccines against other common infectious diseases, including AIDS, tuberculosis, and malaria suggest that the current empirical approach to vaccine design is not effective in eliciting protective immunity in many cases [Koff 2013]. This variability in vaccine effectiveness highlights the need to better understand the fundamental principles of human immune responses, the “rules of immunity”, and how this understanding could be used to develop vaccination strategies that are consistently effective and result in durable immunity.

Recently, several groups have applied high-throughput multi-omics assays to produce a comprehensive systems-level evaluation of vaccine responses, so-called “systems vaccinology” (reviewed in [Raeven 2019, Sharma 2019, Kollmann 2020, Wimmers 2020]. One of the key questions addressed is - can baseline (pre-vaccine) signatures of the immune system predict vaccine responses and differentiate between responders vs. non-responders and, if so, what can these signatures tell us about the mechanisms for eliciting protective immunity [Tsang 2020]. The general concept that specific baseline immune signatures can predict vaccine responses has been explored in large cohort studies in the context of hepatitis B virus (HBV), influenza and malaria vaccines [Warimwe 2013, Tsang 2014, Fourati 2016, HIPC-CHI 2017, Bartholomeus 2018, Qui 2018]. However, the connection between these molecular signatures and the underlying immunological mechanisms remains tenuous. Further, the application of large-scale multi-omics assessment of large vaccination cohorts is cost-prohibitive, raising the question of whether advanced computational and machine learning methods may allow for the discovery of predictive mechanistic signatures in studies with smaller sample sizes [Shannon 2020].

The hepatitis B virus (HBV) vaccine is an ideal platform to explore these questions. First, serum anti-HBV antibody levels, which can be easily measured in participant samples, are a well-established correlate of protection [Jack 1999]. Second, the response to the HBV vaccine is highly variable, providing a broad range of responses, which is useful for identifying correlates and predictors [Tsang 2015]. Third, around 10% of subjects respond with protective antibody titres following a single dose [Schillie 2013]. We recently applied a series of validated multi-omics assays to measure the full range of cellular and molecular components of the immune system, including immune cell composition, DNA methylation, gene expression, protein abundance, and fecal 16S microbiome, to provide an exhaustive picture of the immune response to the HBV vaccine [Ben-Othman 2020]. Multi-omics integrative analysis on these data sets identified a number of candidate baseline predictors of vaccine response using serum antibody titers to the HBV surface antigen following three vaccine doses as the quantitatively-defined endpoint in a relatively small cohort of 15 vaccine recipients [Shannon 2020]. While these candidate predictive signatures could be identified using this systems-level approach in a relatively small cohort, a unifying mechanistic driver did not emerge. Furthermore, multi-omic analysis of whole blood failed to reveal features predictive of the variable protective responses following only a single vaccination dose.

In this report, we sought to determine if a more granular approach, namely single cell RNA sequencing in the context of an integrated multi-omic analysis, could identify the relevant cellular phenotypes and functions associated with vaccine responses. Using machine learning to identify the most discriminative gene expression features for dimensionality reduction to optimize performance of predicitive modeling using canonical correlation analysis (CCA), a truly integrative machine learning approach emerged that helps to overcome small sample sizes through a hypothesis-generation-and-hypothesis-testing-in-orthologous-datasets workflow.

## Methods

Descriptions of all methods not detailed below have recently been published in [Ben-Othman 2020, Shannon 2020].

### Cohort and sampling description

A prospective, observational study (ClinicalTrials.gov: NCT03083158) of immune responses to the HBV vaccine (ENGERIX^®^-B) was undertaken, with recruitment occurring at the Vaccine Evaluation Center (VEC), British Columbia Children’s Hospital Research Institute in Vancouver, Canada. Briefly, a total of 15 eligible individuals aged 44 – 73 were enrolled in the study. One ml (20 µg) of ENGERIX^®^-B vaccine was administered via intramuscular deltoid injection at three different times during the study (Day 0, Day 28 and Day180). HBV titres were measured at screening, Day 28 after the initial vaccine dose (immediately prior to second dose), Day 180 (immediately prior to the third dose), and Day 208 (one month after the final dose). For the purposes of this study, participants were categorized as “responders” if their anti-HBV serum antibody titer was >10 IU/ml at Day 28 after a single dose of HBV vaccine, (a response considered as protective), “marginal responder” if they had detectable anti-HBV serum antibody titer above baseline but <10 IU/ml at Day 28, or “non-responders” if they had no detectable anti-HBV serum antibodies at Day 28. Note that due to funding constraints, only a subset of participants and samples were used for some of the mechanistic assays.

Various omics studies were performed as described [Ben-Othman 2020, Shannon 2020]. Briefly, peripheral whole blood cells were profiled by flow cytometry, genome-wide DNA methylation (Illumina Infinium MethylationEPIC BeadChip), transcript abundance (bulk RNA-Seq), and proteome-wide protein abundance (mass spectrometry) at various time points. Additionally, the bacterial composition (microbiome) of the gut was assessed by 16S rRNA microbiome profiling pre-(Day −14 and Day 0) and post-vaccination (Day 14). Procedures for the collection and processing of PBMC samples for single cell RNA sequencing are described below.

### Single cell RNA sequencing

Four innate immune cell subsets (monocytes, natural killer (NK) cells, myeloid dendritic cells (mDCs), and plasmacytoid dendritic cells (pDCs)), were single cell sorted for RNA sequencing as described [Ben-Othman 2020] (Supplementary Figure 1). Briefly, 1.5 ml blood samples were stained and single cells sorted for each cell populations of interest before performing subsequent single cell RNA sequencing. 20 mM of EDTA (Fisher #BP120-500) was diluted 1:10 in 1.5 ml blood and red blood cells lysed by adding RBC lysis buffer (eBiosciences, cat #00-4333-57) per manufacture’s recommendations. After 10 min at room temperature, PBS was added and the cell suspension separated by centrifugation at 500 × g for 5 min. The cell pellet was resuspended in an antibody mixture diluted in PBS and 0.5% BSA (bovine serum albumin, Sigma Aldrich, cat #A7906) according to the manufacture’s recommendations. APC-eFluor 780 (cat #65-0865-14) viability dye was added to cells prior to staining to sort viable cells. The cell mixture was incubated at room temperature in the dark for 30 min, then washed once in PBS, and resuspend in 3 ml of PBS for immediate single cell sorting into wells of a 96-well microtiter plate chilled on ice using a BD FACS Aria. Innate immune cell subsets were sorted from the APC-eFluor 780-viable cell gate as follows: NK cells (CD45+CD66-CD14-HLA-DR-CD3-CD16+), monocytes (CD45+CD66-CD14+), mDCs (CD45+CD66-CD14-HLA-DR+CD11c+), and pDCs (CD45+CD66-CD14-HLA-DR+CD11c-CD123+). Prior to sorting, 96-well plates were pre-loaded with 2μl lysis buffer (0.2% Triton X-100, (Sigma Aldrich, cat #9002-93-1), 2 Units/μl RNase inhibitor (Applied Biosystems, cat #N8080119), 1:2,000,000 dilution of ERCC spike-in RNAs (Life Technologies, cat #4456740) and centrifuged at 300 x g at 4°C for 1 minute to distribute liquid in the bottom of the well. After sorting, each plate containing the single cell lysates was immediately sealed, frozen on dry ice, and stored at − 80°C.

**Figure 1.**
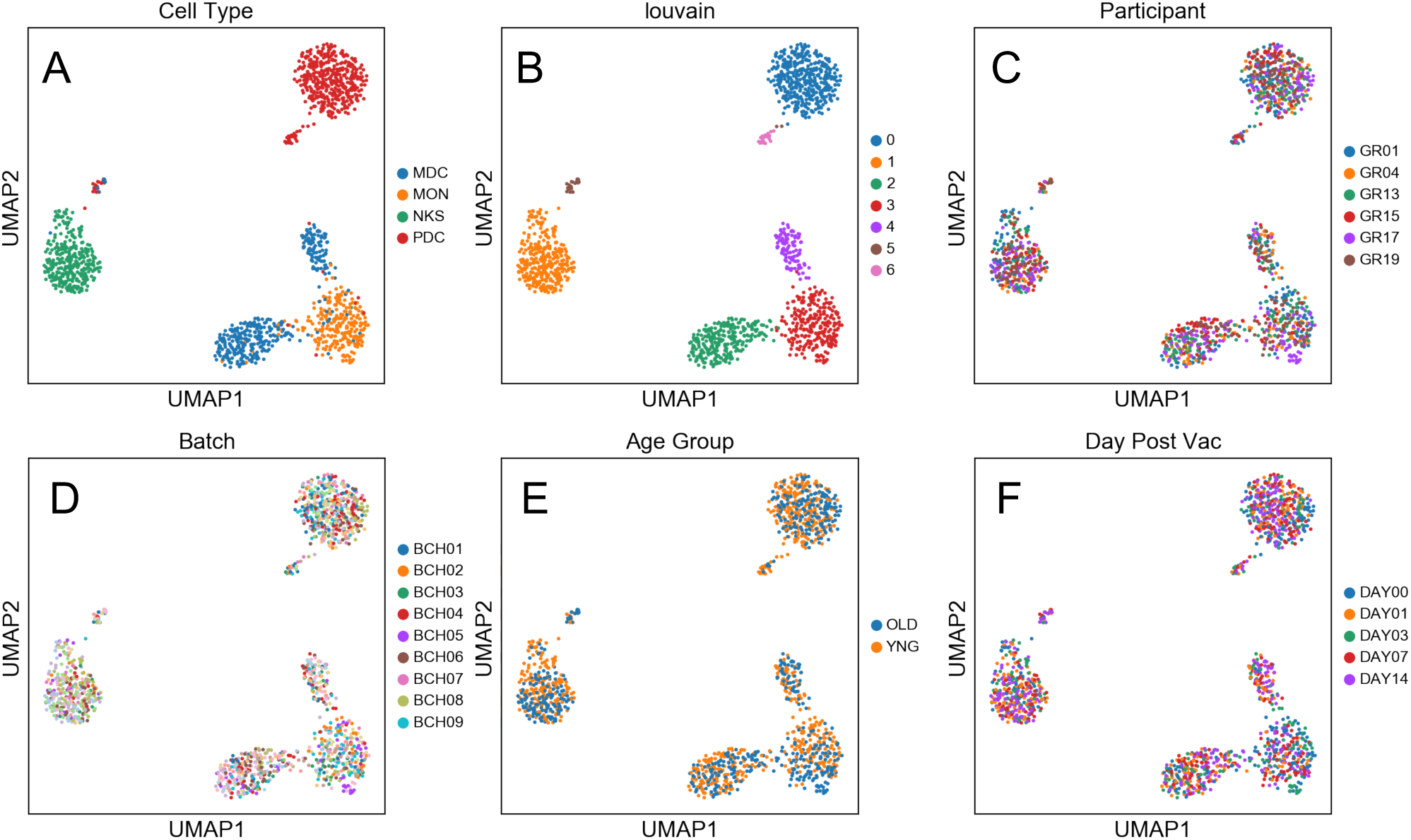
Two distinct mDC subsets are found in blood of participants using scRNAseq. UMAP embedding of transcriptional profiles and Louvain clustering results reveal seven expression clusters from the four sorted innate immune cell populations, including two distinct mDC clusters (Louvain Cluster #2 and #4). Coloring corresponds to FACS-sorted cell type (A), Louvain cluster membership (B), Participant ID (C), sample processing batch (D), age group (E), and sample collection day before or after vaccination (F).

Processing of the frozen 96-well plates containing single cell lysates was performed as previously described [Krishnaswami 2016] with modifications to accommodate an Agilent BioCel automated liquid handling platform [McClean 2013]. Briefly, single cell lysates contained in the 96-well sorted plates were processed in batches of eight plates, with each plate containing wells reserved for 10 pg Universal Human RNA (Clontech Cat#636538) as a positive control, an ERCC-only control, and water as a negative control. Smart-seq2 cDNA synthesis, reverse transcription, and PCR were carried out in a reduced volume (12.5 µL) and with ERCC internal controls spiked-in at a reduced concentration (55 million-fold dilution of the ERCC stock in the first strand cDNA synthesis step). Amplified cDNAs from the eight 96-well plates were consolidated to two 384-well plates and purified with Ampure magnetic particles. A 10-fold diluted portion of each cDNAs was assessed for expression of the human beta-actin (ACTB) housekeeping gene by qPCR for quality control of the amplified cDNAs. A total of 14,592 sample wells were processed through cDNA synthesis and ACTB qPCR on the automated platform.

A cycle threshold (Ct) of ≤35 for ACTB amplification was used as a cutoff for the selection of 3,072 cDNAs (768 per cell type) for library preparation and sequencing. A Star liquid handling platform (Hamilton) was used to consolidate cDNAs selected for Illumina Nextera XT library preps (Illumina cat# FC-131-1096) into 384-well plates. An automated 1/8th Nextera XT reaction was carried out on 125 pg of the selected cDNAs for the Tn5 tagmentation step, with limited 15 cycle PCR followed by AmPure XP (Beckman Coulter Cat# A63881) bead purification. Nextera XT PCR was carried out with a combination of 384 barcode pairs using Nextera XT Index Kit V2 barcode sets A and D (Illumina cat# FC-131-2001 and −2004). Concentrations of the purified Nextera XT reactions were normalized to 1 ng/µL and combined into a 2ng pool of 384 dual-barcoded samples. RNA-seq was carried out with a total of eight 384 barcoded pools loaded across 16 lanes of an Illumina HiSeq 2500 according to manufacturer’s specifications for a total of 3,072 samples sequenced, including controls. A HiSeq SBS V4 250 cycle kit (Illumina cat# FC-401-4003) and a Paired End V4 Cluster Kit (Illumina cat# PE-401-4001) were used for an estimated 2 million reads per sample.

### Sequence data processing

Single cell RNA-seq data was processed according to published methods [Krishnaswami 2016, Aevermann 2017]. Briefly, raw fastq sequencing files were demultiplexing using Illumina barcodes. Sequencing primers and low-quality bases were removed using the Trimmomatic software package [Bolger 2014]. Trimmed reads were then aligned using HISAT [Pertea 2016] in two steps: first to a reference of ERCC sequences, and then to GRCh38 (Ensembl). StringTie [Pertea 2016] was used to assemble the resulting alignments into transcript structures using GENCODE v25 annotation (Ensembl 87; 10-2016) and gene expression values (TPM) estimated. HTSeq-count [Anders 2015] was used to generate raw gene alignment counts.

Quality control analysis was performed using sequencing and laboratory metrics, including average Phred score, percent duplicate reads, and transcript isoform count, to classify cell samples as pass or fail using a Random Forest quality control classification model previously described [Aevermann 2017]. Expression values for the top 2500 genes ranked based on variance from cell samples that passed quality control classification were fed into Scanpy [Wolf 2018] for principal component analysis (PCA) and Uniform Manifold Approximation and Projection (UMAP)-based non-linear dimensionality reduction and visualization [McInnes 2018, Becht 2018]. Unsupervised clustering was performed for the entire dataset, while additional supervised clustering guided by flow cytometry marker panels was performed to investigate within cell type variation. Lastly, cell type marker determination was performed using the Louvain unsupervised clustering results and the NS-Forest algorithm [Aevermann 2018, Aevermann 2020]. The end result of this computational pipeline produced a set of unbiased cell type clusters, a gene expression matrix with the expression levels of genes in individual single cells grouped into cell type clusters, and a set of sensitive and specific marker genes for each cell type cluster for use in downstream quantitative PCR assays and semantic representations [Bakken 2017].

### qRT-PCR

Aliquots (2 µL) of the Smart-seq2 cDNAs from single sorted myeloid dendritic cells (mDCs) were diluted 10-fold in low TE (10mM Tris, 0.1mM EDTA) and 2.5 µL of the diluted cDNAs were subjected to 10 µL Taqman™ qPCR assays for the human beta actin (ACTB) housekeeping gene (ThermoFisher Hs01060665_g1 FAM-MGB) using 5 µL of a 2X PerfeCTa qPCR SuperMix ROX (Quantabio cat# 95050-500) as an initial screen for endogenous gene expression. Thermocycling conditions were completed on a Quantstudio 6 qPCR instrument (Applied Biosystems) using the following thermocycling profile: initial 95°C activation for 2 minutes followed by 45 cycles of 95°C for 10 seconds and 60°C for 30 seconds. Positive reactions −cycle threshold (Ct) of less than 35 for ACTB amplification - were identified and their corresponding cDNAs screened using two additional marker genes selected from the NS-Forest analysis, CDKN1C (ThermoFisher Hs00175938_m1 FAM-MGB) and NDRG2 (ThermoFisher Hs01045114_g1 FAM-MGB), using the same thermocyling conditions for ACTB.

### In vitro mDC stimulation

Pre-vaccination (baseline) blood samples were stimulated *in vitro* (Milieu Interieur) with LPS, poly IC, or SEB with appropriate negative controls and incubated in TruCulture tubes within 15 minutes of blood collection, inserted into a dry block incubator, and maintained at 37°C (±1°C) for 22 hours as described [Ben-Othman 2020, Shannon 2020]. Cell fractions were collected and lysed in Trizol for RNA extraction. cDNA was prepared using the SmartSeq 2 protocol as described above. Quantitative PCR (qPCR) was performed in triplicates for each sample targeting CDNK1C and NDRG2 using ACTB as a housekeeping genes. The data were analyzed using the delta-delta CT method.

### mDC functional assessment

To assess the ability of mDCs to induce T cell activation, 50 ml of whole blood was drawn from healthy adult donors and PBMC isolated as previously described [Ben-Othman 2020]. Cells were stained using cocktails of surface marker antibodies (FCER1A, CD11c, CD1C, CD14, CD3, CD123, CD16A and HLADR) to specifically sort mDC2, mDC4 and T cells (Supplementary Table 1, Supplementary Figure 2). Cell sorting was performed using BD Aria (II 85µm nozzle) in cold chambers. Sorted cells were spun down at 600g for 10 min, resuspended in 1ml PBS, counted and seeded in 96 well plates pre-filled with either LPS (100 ng/ml) or polyI:C (20µg/ml) or medium as a negative control. The autologous T cells were labelled with Cell Trace ™ Oregon Green diluted in PBS according to the manufacturers instructions (Invitrogen # C34555), and then rested in AIM V medium (Gibco Cat# 12055-091) with 2% heat inactivated human AB serum (e.g. Gemini BioProducts) at 37°C for 24 hr. The next day, DC cultures were briefly treated with 20 mM of EDTA to detach adherent cells, all cells harvested, washed three times in complete medium to remove the TLR ligands, and counted. Dendritic cells (mDC2 or mDC4) were then mixed with labelled T cells at a ratio of 1:5 in 125µl complete AIM V medium and incubated at 37°C for 5 days. On day 5, the cells were detached using EDTA, washed and stained with a cocktail of antibodies (Supplementary Table 2) to assess the proliferation of specific T cells using BD LSRII flow cytometer. All flow cytometry data were analyzed using Flowjo version 10 (Flowjo, Ashland, OR).

**Figure 2.**
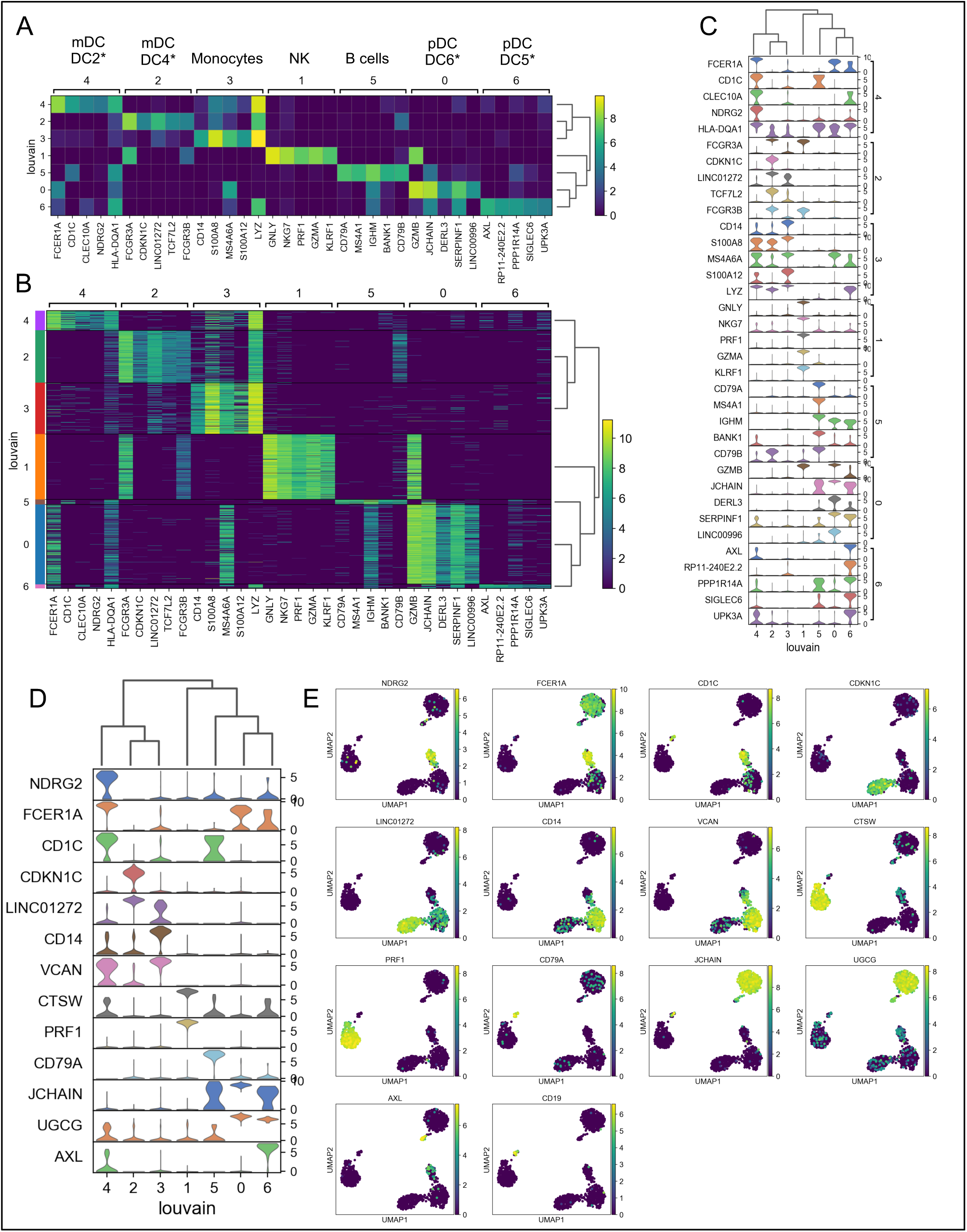
Expression cluster marker genes. A) The top five marker genes for each cluster was determined by logistic regression. Median expression of marker genes in cells within each cluster is shown. *Dendritic cell types identified in Villani et al. [Villani 2017]. B) Expression of logistic regression marker genes in each individual cell within each cluster. C) Violin plots showing logistic regression marker gene expression distributions. D) Violin plots showing gene expression distributions for the minimum set of necessary and sufficient marker genes as determined using the NS-Forest algorithm. E) Expression of NS-Forest marker genes in UMAP Louvain clusters.

### Diablo supervised sGCCA

DIABLO, part of the mixOmics framework, is a data-driven, hypothesis-free multi-omics integration approach that has been successfully applied, by us and others, to derive novel, robust biomarkers, and increase our understanding of the molecular regulatory mechanisms that underlie health and disease [Langenberg 2020, Lee 2019, Rohart 2017, Singh 2019]. DIABLO extends sparse Generalized Canonical Correlation Analysis (sGCCA) into a supervised multi-omics data integration framework [Le Cao, 2011, Tenenhaus 2014]. DIABLO performs multivariate dimensionality reduction and selects correlated variables from several datasets by maximizing the covariance between linear combinations of variables (latent component), across both multi-omics datasets (blocks) and an outcome (response) variable, in this case anti-HBV serum antibody titers. The data are then projected into a smaller dimensional subspace spanned by the latent components for classification. Here we used DIABLO to identify baseline (pre-vaccination) predictors of vaccine response (anti-HBV IgG level measured at the final follow-up visit, Day 208), from multi-omics profiles in an integrative fashion.

## Results

### mDC subsets with distinct gene expression

Single cell RNA sequencing (scRNAseq) of innate immune cell subsets sorted from whole blood was used to define their transcriptional phenotypes with relationship to HBV vaccination responses. In order to ensure the capture of any distinct phenotype that might predict response, blood samples were collected on Day 0 pre-vaccination and Day 1, 3, 7, and 14 post-vaccination. The response to the vaccine (according to HBV-specific antibody titre levels at 1 month) was a criteria to differentiate responders from non-reponders based on the well established correlate of protection of 10mIU/ml as previously described [Shannon 2020]. Using those criteria, 2 responders (GR01, GR04), 1 marginal responder (GR15), and 3 non-responders (GR13, GR17, GR19) to a single dose of vaccine were chosen for analysis. Single monocytes (MON), myeloid dendritic cells (mDC), plasmacytoid dendritic cells (pDC), and natural killer cells (NK) were sorted into microtiter plate wells and single cell cDNAs showing positive ACTB expression by qPCR analyzed by scRNAseq.

Each of the four major innate immune cell subsets were well segregated in the UMAP plot (Figure 1A). In addition, lower abundance outlier clusters we also detected for the mDC and pDC sorted populations, indicating some level of subtype heterogeneity. Unsupervised clustering produced seven distinct transcriptome clusters (Figure 1B), including the lower abundance mDC and pDC outliers. No obvious cluster-specific enrichment of cells from individual participants, processing batch, age group or sample collection date was observed (Figures 1C – F).

Each unsupervised cell cluster showed distinct differential gene expression patterns identified using both logistic regression (Figure 2A-C) and NS-Forest-based marker gene selection (Figure 2D-E and Supplemental Table 3). The main mDC subset (Louvain cluster #2) appeared to exclusively express the p57 kip2 cyclin-dependent kinase inhibitor gene CDKN1C and expressed relatively high levels of LINC01272 in comparison with other innate cell subsets (Figure 2A-E). These cells also expressed high levels of the Fc gamma receptor gene FCGR3A (Figure 2A-C). In contrast, the outlier mDC cluster (Louvain cluster #4) exclusively expressed the n-myc regulated gene NDRG2 and expressed relatively high levels of the Fc epsilon receptor gene FCER1A, the MHC class II gene HLA-DQA1 (Figure 2A-E), and other MHC class II genes (not shown). The high-level expression of MHC class II genes suggests that the outlier mDC subset is activated, whereas the expression of the p57 kip2 CDK inhibitor suggests that the main mDC subset is resting. Expression of FCGR3A by the main mDC cluster and FCER1A by the outlier mDC cluster suggests that these two subsets correspond to the DC4 and DC2 dendritic cell types defined previously [Villani 2017] and will be referred to as NDRG2-expressing mDC2 and CDKN1C-expressing mDC4.

In order to determine if the NDRG2-expressing mDC2 and CDKN1C-expressing mDC4 phenotypes had been observed in previous studies, the NDRG2 and CDKN1 marker genes were used to search for expression modules in the MSigDB database. Four MSigDB modules included both of these marker genes:

- GSE17721_0.5H_VS_24H_POLYIC_BMDC_UP,
- GSE17721_0.5H_VS_8H_POLYIC_BMDC_DN,
- GSE17721_CTRL_VS_POLYIC_12H_BMDC_DN,
- GSE17721_POLYIC_VS_PAM3CSK4_12H_BMDC_UP),

which were all derived from dendritic cells stimulated with TLR3 agonists.

### Relative abundance of mDC subsets correlate with vaccine response

Exclusive expression of NDRG2 in mDC2 and CDKN1 in mDC4 suggested that these two markers could be used to distinguish these mDC subsets. Indeed, qPCR amplification showed mutually exclusive expression of these two genes in the sorted mDCs used for scRNAseq (Figure 3B). qPCR for NDRG2 and CDKN1C was used to identify and quantify these two mDC subsets in a larger cohort of ten participants across the entire time course of the study (Supplementary Tables 4 & 5). The relative proportions of NDRG2-expressing mDC2/CDKN1C-expressing mDC4 were found to be dynamic and vary between individuals (Figure 3B). Interestingly, a relatively high ratio of NDRG2-expressing mDC2/CDKN1C-expressing mDC4 was found in the two single-dose responders (GR01 and GR04) at baseline Day 0 (Supplementary Table 5). Indeed, the average NDRG2-expressing mDC2/CDKN1C-expressing mDC4 ratio in responders on Day 1 was 3.13 and in non-responders was 0.46. Interestingly, while the ratio of NDRG2-expressing mDC2/CDKN1C-expressing mDC4 dropped dramatically following vaccination of the two responders, the ratio was relatively static or increased in the non-responders (Figure 3A). While these finding in-and-of themselves are not adequately powered to draw solid conclusions regarding correlation due to the small sample size of this pilot, it did produce a hypothesis that could be explored in orthologous data, namely that the relative proportions of these two distinct mDC subsets might be predictive of HBV vaccine responses.

**Figure 3.**
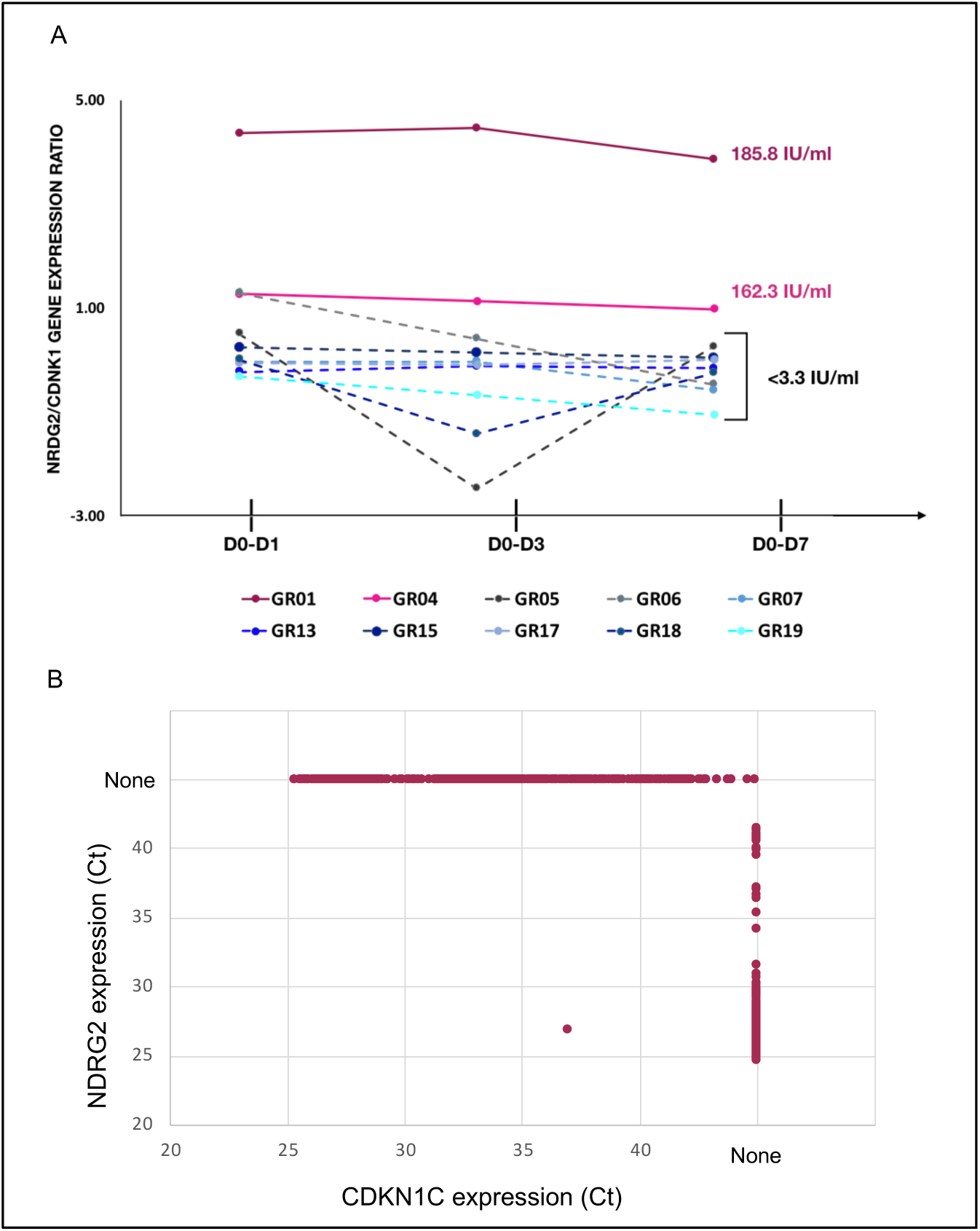
Relative proportion of mDC subsets expressing NDRG2 and CDNK1 are predictive of HBV vaccine response. A) Single myeloid dendritic cells were sorted from blood collected prior to HBV vaccination (D0) and 1 day (D1), 3 days (D3) and 7 days (D7) post vaccination. Following cDNA preparation, the expression of NDRG2 (mDC2 expressing gene) and CDNK1 (mDC4 expressing genes) mRNAs were quantified by qPCR. The graph shows the change in the relative proportion of NDRG2-expressing mDC2s/CDKN1C-expressing mDC4s at each time point compared to D0 per study participants. Solid lines show the HBV early responders (with anti-HBV titres higher than 3.3 IU/ml at Day 28 after first dose and titers indicated next to the lines). Dotted lines show the participants with no measurable response (less than 3.3 IU/ml HBV titers) after the first dose of vaccine. Each line shows values of individual participants; the Y axis values were log transformed. Raw data is provided in Supplemental Figure 3. B) Ct values for qPCR reactions measuring expression of NDRG2 and CDKN1C for 964 single mDC cells expressing at least one marker are plotted, showing mutually-exclusive expression of NDRG2 and CDKN1C in sorted mDCs. None indicates no amplification.

Given that NDRG2-expressing mDC2 and CDKN1C-expressing mDC4 marker genes in the MSigDB database had been identified as malleable to polyI:C stimulation, we further explored how TLR3 activation might lead to differential responses in these DC subsets, we examined the changes in expression of CDKN1C and NDRG2 in whole blood collected from participants prior to HBV vaccination and stimulated with a TLR3 agonist. Interestingly, the two participants that showed the highest Ab responses to the first vaccine dose – GR01 and GR04 – showed preferential up-regulation of NDRG2 in their blood cells suggesting they were predisposed to respond with a more activating mDC2-like phenotype, whereas cells from non-responding participants showed preferential up-regulation of CDKN1C suggesting they were predisposed to a more resting/inhibitory mDC4-like phenotype (Supplementary Figure 3).

### mDC subsets differ in their functional predisposition

To explore if there are any functional differences between these mDC subsets, NDRG2-expressing mDC2 and CDKN1C-expressing mDC4 subsets were sorted and incubated with sorted T cells from the same (autologous) donor. Furthermore, sorted DCs were also pre-stimulated with TLR3 (pIC) or TLR4 (LPS) agonists prior to co-culture with autologous T cells to assessed the impact of TLR pre-stimulation on T cell proliferation as a proxy read out for immune activation as described [Villani 2017]. The positive control, i.e. proliferation of SEB stimulated T cells, was not impacted by co-culture of either unstimulated or stimulated mDC2 or mDC4 (data not shown). When unstimulated CDKN1C-expressing mDC4 cells were incubated with unstimulated autologous T cells, T cell proliferation was induced (Figure 4A & B). However, if CDKN1C-expressing mDC4 cells were first stimulated with pIC or LPS, this baseline T cell proliferation was inhibited 3.9 fold (mean n = 4) for both CD4 and CD8 T cells (paired t-test p = 0.030 and 0.036 respectively). In contrast, NDRG2-expressing mDC2 cells did not induce T cell proliferation, with or without TLR stimulation (data not shown). These results suggest that TLR activation of CDKN1C-expressing mDC4 cells suppresses their tonic T cell activating ability, while TLR3 or TLR4 stimulation of mDC2 does not.

**Figure 4.**
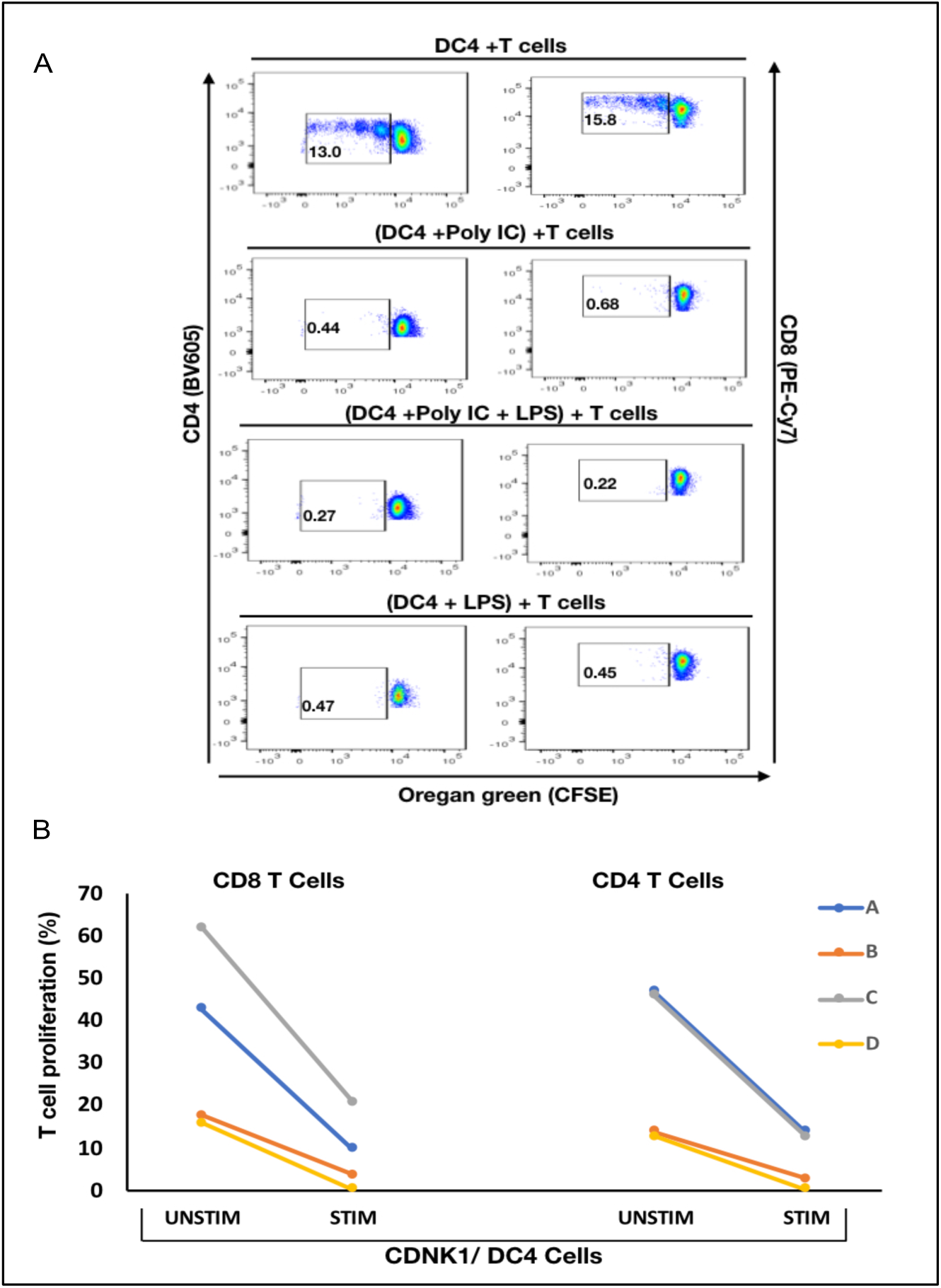
Stimulation of CDKN1C-expressing mDC4 cells with TLR agonists suppresses T cell activation function. A) CDKN1/DC4 cells were sorted as viable, singlets, CD14-, CD19-, CD3-, HLA-DR^int-to-high^, CD11c+, CD16+, stimulated with polyIC and/or LPS and mixed with autologous T cells labelled with Oregan Green (OG) from the same patient. (Note that CD16 is encoded by FCGR3A expressed in CDKN1C-expressing mDC4.) Following 5 days, samples were analyzed by flow cytometry. Proliferating T cells (lower OG staining) were gated. The pseudocolor dot plot is representative of 4 different experiments. B) Percent of T cells proliferating using sorted cells from four different individuals with or without stimulated. Line graphs of a composite of T cells proliferation assay data (CD4+ and CD8+) (n=4). Proliferation was assessed using OG staining after co-culture of autologous T cells with CDKN1C-expressing mDC4 cells for 5 days induced by either unstimulated or pre-stimulated with the TLR3 agonist polyIC. Each coloured line (A, B, C and D) represents proliferation data from one of four different subjects.

### Machine learning model using mDC TLR3 feature selection predicts serum antibody responses

In our previous study, we found that multi-omics data could be used to produce predictive models of antibody titers based on the supervised sparse generalized canonical correlation analysis implemented in the Diablo algorithm [Shannon 2020], even given the relatively large feature space provided by the transcriptomic and CpG methylation data (Figure 5A). Based on the scRNAseq and functional studies described above, we hypothesized that if the relative abundance of the NDRG2-expressing mDC2 and CDKN1C-expressing mDC4 subsets were indeed mechanistically linked to vaccine responses, selecting features specific to these cell subsets might produce better predictive models. Using just the bulk gene expression and DNA methylation data associated with the MSigDB marker genes derived from dendritic cells stimulated with TLR3 agonists to build the Diablo predictive model, a significant improvement in model performance in training was obtained (Figure 5B). For example, the Spearman’s rank correlation of the gene expression model improved from 0.62 to 0.87 and the median error improved from 15.66% to 7.23%. Similar improvements were observed with the model produced using the selected CpG dna methylation features. And while cross-validation did not show a significant improvement, likely due to the small sample size, we went from ~50,000 transcripts and ~800,000 CpG sites to only a few hundred transcripts and a few thousand CpG sites demonstrating a substantial enrichment of informative features. These results indicate that feature selection based on TLR3-induced dendritic cell genes produced machine learning models that provided better prediction of serum antibody responses, suggesting that the relative contributions of the NDRG2-expressing mDC2 and CDKN1C-expressing mDC4 subsets influence this vaccine response.

**Figure 5.**
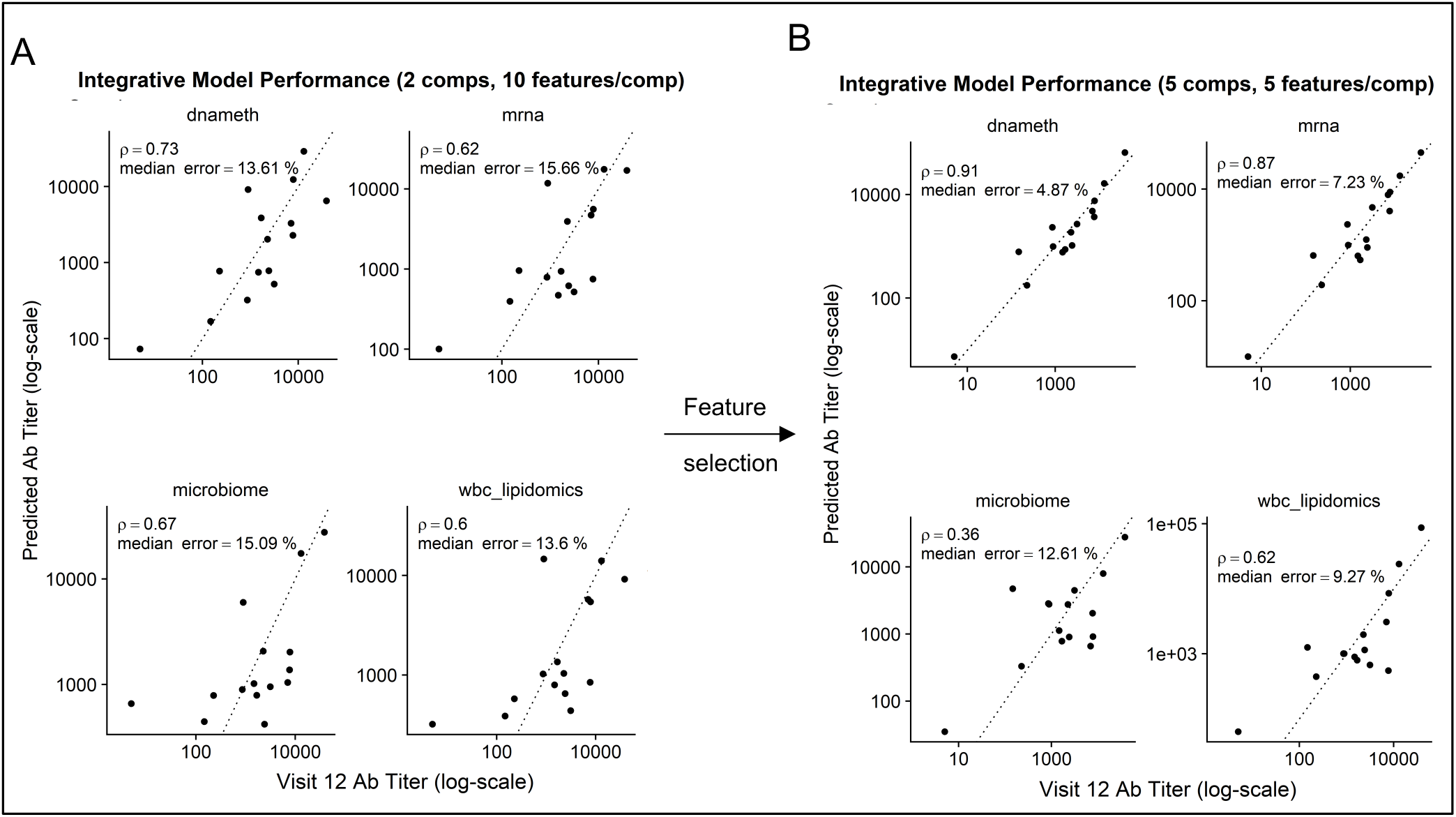
Performance of Diablo models in predicting serum antibody responses to HBV vaccine. Actual (x-axis) vs. predicted (y-axis) vs. antibody titers at Visit 12 (after third dose) in models produced using different assay platforms are visualized. Dotted line is the identity line representing perfect prediction. Rho is Spearman’s rank correlation when comparing actual Ab titers to predicted Ab titers. A) Diablo models with 2 components and 10 features/component were built using all available assay variables. B) Diablo models with 5 components and 5 features/component were built using selected variables related to 735 dendritic cell TLR3-response genes in the DNA methylation (dnameth) and bulk transcriptomic (mrna) data. CpG sites were assigned to dendritic cell TLR3-response genes as described previously [Price, 2013, Pidsley 2016]. Note that although the microbiome and wbc-lipidomics data are identical in the two sets, the models they produce (features retained and their coefficients) in the generalized canonical correlation framework are different due to different correlation characterisics with the different dnameth and mrna models. In both cases, the number of components and number of features/component were selected to maximize model performance.

## Discussion

In a relatively small cohort of HBV vaccine recipients, we identified two distinct mDC subsets using single cell transcriptomics analysis, the ratio of which at baseline (i.e. before vaccination) correlated with vaccine response to a single dose of the HBV vaccine. These two mDC subsets were distinguishable by the differential expression of a number of genes that allowed for their putative matching to dendritic cell subsets identified previously [Villani 2017], designated here as NDRG2-expressing mDC2 and CDKN1C-expressing mDC4.

Three pieces of evidence suggest that the relative contributions of these dendritic cell subsets to the immune state at baseline prior to vaccination influences vaccine responses. First, the two individuals who generated serum antibody responses to a single dose of HVB vaccine had a high ratio of NDRG2-expressing mDC2/CDKN1C-expressing mDC4 in their peripheral blood prior to vaccination. Second, whole blood from these responding individuals showed preferential upregulation of NDRG2 when stimulated with a TLR3 agonist. Third, machine learning-derived predictive models built using genes differentially expressed in dendritic cells stimulated with TLR3 agonists outperformed models built without this feature selection step. Thus, the dispositional state of these dendritic cell subsets appears to provide a baseline predictor of HBV vaccine responses.

The mechanism of how NDRG2-expressing mDC2 and CDKN1C-expressing mDC4 impact the vaccine response to the HBV vaccine is not yet clear. But, interestingly, *ex vivo* CDKN1C-expressing mDC4 cells were able to induce autologous CD4 and CD8 T cell proliferation yet mDC2 did not; and TLR3 or TLR4 stimulation of the mDC4 subset inhibited this T cell stimulation. Such a high state of functional plasticity in the mDC4 subset, and the longitudinal variation over time in the ratio of NDRG2-expressing mDC2 and CDKN1C-expressing mDC4 in whole peripheral blood may at least partially explain the variability in immune responses to the HBV vaccine.

Several other groups have recently explored the identification of baseline predictors of vaccine responses using multi-omics assays with results that differ from those reported here. Tsang et al. used multi-omics assays to explore responses to seasonal influeza vaccine in healthy adults [Tsang 2014]. While neither Day 0 gene expression nor pathway analysis alone was predictive of vaccine responses in their study, twelve cell populations assessed by flow cytometry, including memory, naive, and transitional B cells, CD4 effector memory T cells, IFNa+ myeloid dendritic cells (mDC), and several activated T cell populations correlated with mean fold change in antibody titers. However, the responses in these healthy influenza vaccine cohorts likely represent a recall response to antigenically-similar prior exposure as opposed to the primary HBV response in our study. Fourati et al. constructed a naive Bayes classifier based on the top 15 differentially-expressed genes between PBMCs from responders and poor-responders to the HBV vaccines, which included B cell markers (e.g., CD20, IGHG1), downstream targets of B-cell receptor signalling (e.g., BANK1) and molecules known to have functional interactions with IgG (e.g., C1, FCGR3B), with a predictive accuracy of ~63% [Fourati 2016]. However, the use of bulk transcriptomic analysis of PBMCs may have obscured the contribution of the minor dendritic cell component evaluated in our scRNAseq assays, emphasizing the value of using scRNAseq to assess the contribution of rare cell subsets. Bartholomeus et al. found that the GRN and IFITM1 genes were significantly downregulated in responders while upregulated in non-responders by whole blood transcriptomics analysis, and absolute granulocytes numbers were significantly higher in non-responders at Day 0 prior to vaccination with Engerix-B [Bartholomeus 2018]. However, the role of the dendritic cell component was not evaluated.

Our results suggest that the participants in our vaccine study existed in one of two different dendritic cell dispositional states at baseline – an NDRG2-mDC2 state in which the vaccine elicited an early antibody response or a CDKN1C-mDC4 state in which the vaccine response may have been actively suppressed. While the possibility that challenge with a foreign antigen (e.g. a vaccine) would be immunosuppressive seems counterintuitive, a healthy immune system has to strike a delicate balance between responsiveness and non-responsiveness under many circumstances. Furthermore, ample data now supports the conclusion that for some vaccines, including ENGERIX^®^-B, reducing general immune activation and inflammation may in fact increase the antigen-specific response to the vaccine [Alter 2015, Fourati 2016].

From a biochemical perspective, foreign antigens are not that different from self-antigens. In order to avoid autoimmunity, the immune system must carefully assess whether an antigen is truly foreign or not. Evidence suggests that the setting in which the naïve immune system experiences antigen may play an important role and is only activated when some type of tissue injury signal accompanies antigen exposure, the so-called “danger hypothesis” [Matzinger 1994]. In the absence of this danger signal, activation of adaptive immune cells with Signal 1 from antigen without Signal 2 from antigen presenting cell help would be tolerated and not result in activation. And even when the immune system responds to a truly foreign antigen, an overexuberant response (e.g., the cytokine storm) can do more harm than good. Thus, in addition to mechanisms designed to activate an immune response, the immune system has also developed mechanisms for dampening the response, with the regulatory T cell being a classic example. Perhaps the CDKN1C-mDC4 cell is an example of a suppressive type of regulatory dendritic cell. The extent to which these CDKN1C-mDC4s may be similar to the myeloid-derived suppressor cells (MDSCs) observed in some pathological conditions, such as inflammation, chronic infection or cancer [Palucka 2011, Sendo 2018, Dorhoi 2018, Karin 2020] remains to be determined.

So, what are the implications of this hypothesis for improving vaccination outcomes? We found that the NDRG2-mDC2/CDKN1C-mDC4 ratio differed between individuals as well as within the same individual over time. This suggests that perhaps the dendritic cell dispositional state could be modulated to establish an activatable predisposition prior to vaccination. Since adjuvant effects appear to function through the innate immune system, perhaps prior exposure to adjuvant *before* antigen could establish the appropriate activatable predisposition. While there is some evidence that preconditioning injection sites with TLR agonists can enhance dendritic cell migration [Tripp 2010] and protection against pathogen infection [Lipford 2000] in animal models, to our knowledge this has never been formally assessed in humans [Tsang 2020]. This possibility could lead to a more precision-medicine approach for vaccines by determining at baseline who will respond well or not to specific vaccines or who might need just a single dose [Poland 2018]. This approach could readily follow the model of point-of-care testing currently used in infectious disease settings (e.g. [Pennisis 2021]). In fact, the field of vaccinology already does assign different vaccines based on individual characteristics, e.g. different flu vaccines are given to different people based on age.

Finally, we described a novel machine learning approach to multi-omics data integration. The results reported by Shannon et al. [Shannon 2020] suggested that because different sources of background noise and technical confounders would contribute to the results from different omics assays, focusing on the consensus information related to outcome using the canonical correlation analysis approach to multi-omics data integration could reduce overfitting and result in more robust and generalizable models [Lee 2019, Singh 2019], even in studies where p>>>n. However, the number of parameter features available in these systems biology studies makes it impossible to complete an exhaustive search of all available parameter combinations and makes the L1 penalized or LASSO regression implemented in Diablo ineffective at mitigating the effects of noisy features [Debashis 2008]. Using single cell RNA sequencing in this study, we were able to identify relatively rare dendritic cell populations whose abundance and activation disposition appear to correlate with vaccine responses. By using this finding to guide feature selection for those genes expressed in these dendritic cell subsets or those CpG sites that are involved in establishing cell type identity, the performance of the vaccine response predictive models was dramatically improved, demonstrating the value of directed feature selection prior to machine learning model production to further circumvent the p>>>n problem.

In conclusion, the machine learning approaches for informative feature selection based on NS-Forest and multi-omics data integration based on supervised canonical correlation analysis not only produced a predictive model of vaccine response but also revealed the possible cellular mechanisms responsible. These results suggest that vaccine recipients exist as different dispositional endotypes that dictate their response to vaccination. With a hypothetical mechanism in hand, developing strategies to adjust these dispositional endotypes in preference of dendritic cell activation rather than suppression could lead to development of more effective precision vaccination strategies to achieve protective immunity from single vaccine doses, which are of critical importance in resource-limited settings.

## Data Availability

The data will be made available after verifying that we have appropriate consent/permission to submit the data to a public archive.

## Acknowledgements

This work was supported by the Human Vaccines Project. Additional funding from the Canadian Institutes for Health Research FDN-154287 to REWH is gratefully acknowledged. REWH holds a Canada Research Chair and a UBC Killam Professorship. DD acknowledges support from the Milieu Interieur Consortium.

## Notes

### Competing Interest Statement

The authors have declared no competing interest.

### Clinical Trial

NCT03083158

### Author Declarations

This project was approved by the University of British Columbia Childrens & Womens Research Ethics Board (Ref: H17-00175)

## Literature cited

Aevermann B, McCorrison J, Venepally P, Hodge R, Bakken T, Miller J, Novotny M, Tran DN, Diezfuertes F, Christiansen L, Zhang F, Steemers F, Lasken RS, Lein ED, Schork N, Scheuermann RH. Production of a Preliminary Quality Control Pipeline for Single Nuclei Rna-Seq and Its Application in the Analysis of Cell Type Diversity of Post-Mortem Human Brain Neocortex. Pac Symp Biocomput. 2017;22:564–575. doi: 10.1142/9789813207813_0052. PMID: 27897007; PMCID: PMC5338304.

Aevermann BD, Novotny M, Bakken T, Miller JA, Diehl AD, Osumi-Sutherland D, Lasken RS, Lein ES, Scheuermann RH. Cell type discovery using single-cell transcriptomics: implications for ontological representation. Hum Mol Genet. 2018 May 1;27(R1):R40–R47. doi: 10.1093/hmg/ddy100. PMID: 29590361; PMCID: PMC5946857.

Alter G, Sekaly RP. Beyond adjuvants: Antagonizing inflammation to enhance vaccine immunity. Vaccine. 2015 Jun 8;33 Suppl 2:B55–9. doi: 10.1016/j.vaccine.2015.03.058. PMID: 26022570.

Anders S, Pyl PT, Huber W. HTSeq--a Python framework to work with high-throughput sequencing data. Bioinformatics. 2015 Jan 15;31(2):166–9. doi: 10.1093/bioinformatics/btu638. Epub 2014 Sep 25. PMID: 25260700; PMCID: PMC4287950.

Bakken T, Cowell L, Aevermann BD, Novotny M, Hodge R, Miller JA, Lee A, Chang I, McCorrison J, Pulendran B, Qian Y, Schork NJ, Lasken RS, Lein ES, Scheuermann RH. Cell type discovery and representation in the era of high-content single cell phenotyping. BMC Bioinformatics. 2017 Dec 21;v18(Suppl 17):559. doi: 10.1186/s12859-017-1977-1. PMID: 29322913; PMCID: PMC5763450.

Bartholomeus E, De Neuter N, Meysman P, Suls A, Keersmaekers N, Elias G, Jansens H, Hens N, Smits E, Van Tendeloo V, Beutels P, Van Damme P, Ogunjimi B, Laukens K, Mortier G. Transcriptome profiling in blood before and after hepatitis B vaccination shows significant differences in gene expression between responders and non-responders. Vaccine. 2018 Oct 8;36(42):6282–6289. doi: 10.1016/j.vaccine.2018.09.001. Epub 2018 Sep 8. PMID: 30205979.

Becht E, McInnes L, Healy J, Dutertre CA, Kwok IWH, Ng LG, Ginhoux F, Newell EW. Dimensionality reduction for visualizing single-cell data using UMAP. Nat Biotechnol. 2018 Dec 3. doi: 10.1038/nbt.4314. Epub ahead of print. PMID: 30531897.

Ben-Othman R, Cai B, Liu AC, Varankovich N, He D, Blimkie TM, Lee AH, Gill EE, Novotny M, Aevermann B, Drissler S, Shannon CP, McCann S, Marty K, Bjornson G, Edgar RD, Lin DTS, Gladish N, Maclsaac J, Amenyogbe N, Chan Q, Llibre A, Collin J, Landais E, Le K, Reiss SM, Koff WC, Havenar-Daughton C, Heran M, Sangha B, Walt D, Krajden M, Crotty S, Sok D, Briney B, Burton DR, Duffy D, Foster LJ, Mohn WW, Kobor MS, Tebbutt SJ, Brinkman RR, Scheuermann RH, Hancock REW, Kollmann TR, Sadarangani M. Systems Biology Methods Applied to Blood and Tissue for a Comprehensive Analysis of Immune Response to Hepatitis B Vaccine in Adults. Front Immunol. 2020 Nov 4;11:580373. doi: 10.3389/fimmu.2020.580373. PMID: 33250895; PMCID: PMC7672042.

Bolger AM, Lohse M, Usadel B. Trimmomatic: a flexible trimmer for Illumina sequence data. Bioinformatics. 2014 Aug 1;30(15):2114–20. doi: 10.1093/bioinformatics/btu170. Epub 2014 Apr 1. PMID: 24695404; PMCID: PMC4103590.

Debashis P, Bair E, Hastie T, Tibshirani R. “Preconditioning” for feature selection and regression in high-dimensional problems. The Annals of Statistics. 2008; 36(4):1595–1618. doi: 10.1214/009053607000000578.

Dorhoi A, Du Plessis N. Monocytic Myeloid-Derived Suppressor Cells in Chronic Infections. Front Immunol. 2018 Jan 4;8:1895. doi: 10.3389/fimmu.2017.01895. PMID: 29354120; PMCID: PMC5758551.

Fourati S, Cristescu R, Loboda A, Talla A, Filali A, Railkar R, Schaeffer AK, Favre D, Gagnon D, Peretz Y, Wang IM, Beals CR, Casimiro DR, Carayannopoulos LN, Sékaly RP. Pre-vaccination inflammation and B-cell signalling predict age-related hyporesponse to hepatitis B vaccination. Nat Commun. 2016 Jan 8;7:10369. doi: 10.1038/ncomms10369. PMID: 26742691; PMCID: PMC4729923.

HIPC-CHI Signatures Project Team; HIPC-I Consortium. Multicohort analysis reveals baseline transcriptional predictors of influenza vaccination responses. Sci Immunol. 2017 Aug 25;2(14):eaal4656. doi: 10.1126/sciimmunol.aal4656. PMID: 28842433; PMCID: PMC5800877.

Jack AD, Hall AJ, Maine N, Mendy M, Whittle HC. What level of hepatitis B antibody is protective? J Infect Dis. 1999 Feb;179(2):489–92. doi: 10.1086/314578. PMID: 9878036.

Karin N. The Development and Homing of Myeloid-Derived Suppressor Cells: From a Two-Stage Model to a Multistep Narrative. Front Immunol. 2020 Oct 26;11:557586. doi: 10.3389/fimmu.2020.557586. PMID: 33193327; PMCID: PMC7649122.

Koff WC, Burton DR, Johnson PR, Walker BD, King CR, Nabel GJ, Ahmed R, Bhan MK, Plotkin SA. Accelerating next-generation vaccine development for global disease prevention. Science. 2013 May 31;340(6136):1232910. doi: 10.1126/science.1232910. PMID: 23723240; PMCID: PMC4026248.

Kollmann TR, Marchant A, Way SS. Vaccination strategies to enhance immunity in neonates. Science. 2020 May 8;368(6491):612–615. doi: 10.1126/science.aaz9447. PMID: 32381718; PMCID: PMC7734703.

Krishnaswami SR, Grindberg RV, Novotny M, Venepally P, Lacar B, Bhutani K, Linker SB, Pham S, Erwin JA, Miller JA, Hodge R, McCarthy JK, Kelder M, McCorrison J, Aevermann BD, Fuertes FD, Scheuermann RH, Lee J, Lein ES, Schork N, McConnell MJ, Gage FH, Lasken RS. Using single nuclei for RNA-seq to capture the transcriptome of postmortem neurons. Nat Protoc. 2016 Mar;11(3):499–524. doi: 10.1038/nprot.2016.015. Epub 2016 Feb 18. PMID: 26890679; PMCID: PMC4941947.

Langenberg MCC, Hoogerwerf MA, Koopman JPR, Janse JJ, Kos-van Oosterhoud J, Feijt C, Jochems SP, de Dood CJ, van Schuijlenburg R, Ozir-Fazalalikhan A, Manurung MD, Sartono E, van der Beek MT, Winkel BMF, Verbeek-Menken PH, Stam KA, van Leeuwen FWB, Meij P, van Diepen A, van Lieshout L, van Dam GJ, Corstjens PLAM, Hokke CH, Yazdanbakhsh M, Visser LG, Roestenberg M. A controlled human Schistosoma mansoni infection model to advance novel drugs, vaccines and diagnostics. Nat Med. 2020 Mar;26(3):326–332. doi: 10.1038/s41591-020-0759-x. Epub 2020 Feb 17. PMID: 32066978.

Lê Cao KA, Boitard S, Besse P. Sparse PLS discriminant analysis: biologically relevant feature selection and graphical displays for multiclass problems. BMC Bioinformatics. 2011 Jun 22;12:253. doi: 10.1186/1471-2105-12-253. PMID: 21693065; PMCID: PMC3133555.

Lee AH, Shannon CP, Amenyogbe N, Bennike TB, Diray-Arce J, Idoko OT, Gill EE, Ben-Othman R, Pomat WS, van Haren SD, Cao KL, Cox M, Darboe A, Falsafi R, Ferrari D, Harbeson DJ, He D, Bing C, Hinshaw SJ, Ndure J, Njie-Jobe J, Pettengill MA, Richmond PC, Ford R, Saleu G, Masiria G, Matlam JP, Kirarock W, Roberts E, Malek M, Sanchez-Schmitz G, Singh A, Angelidou A, Smolen KK; EPIC Consortium, Brinkman RR, Ozonoff A, Hancock REW, van den Biggelaar AHJ, Steen H, Tebbutt SJ, Kampmann B, Levy O, Kollmann TR. Dynamic molecular changes during the first week of human life follow a robust developmental trajectory. Nat Commun. 2019 Mar 12;10(1):1092. doi: 10.1038/s41467-019-08794-x. PMID: 30862783; PMCID: PMC6414553.

Lipford GB, Sparwasser T, Zimmermann S, Heeg K, Wagner H. CpG-DNA-mediated transient lymphadenopathy is associated with a state of Th1 predisposition to antigen-driven responses. J Immunol. 2000 Aug 1;165(3):1228–35. doi: 10.4049/jimmunol.165.3.1228. PMID: 10903720.

Matzinger P. Tolerance, danger, and the extended family. Annu Rev Immunol. 1994;12:991–1045. doi: 10.1146/annurev.iy.12.040194.005015. PMID: 8011301.

McInnes L, Healy J, Melville J. UMAP: uniform manifold approximation and projection for dimension reduction. arXiv 2018. http://arxiv.org/abs/1802.03426.

McLean JS, Lombardo MJ, Ziegler MG, Novotny M, Yee-Greenbaum J, Badger JH, Tesler G, Nurk S, Lesin V, Brami D, Hall AP, Edlund A, Allen LZ, Durkin S, Reed S, Torriani F, Nealson KH, Pevzner PA, Friedman R, Venter JC, Lasken RS. Genome of the pathogen Porphyromonas gingivalis recovered from a biofilm in a hospital sink using a high-throughput single-cell genomics platform. Genome Res. 2013 May;23(5):867–77. doi: 10.1101/gr.150433.112. Epub 2013 Apr 5. PMID: 23564253; PMCID: PMC3638142.

Palucka K, Ueno H, Fay J, Banchereau J. Dendritic cells and immunity against cancer. J Intern Med. 2011 Jan;269(1):64–73. doi: 10.1111/j.1365-2796.2010.02317.x. PMID: 21158979; PMCID: PMC3023888.

Pennisi I, Rodriguez-Manzano J, Moniri A, Kaforou M, Herberg JA, Levin M, Georgiou P. Translation of a Host Blood RNA Signature Distinguishing Bacterial From Viral Infection Into a Platform Suitable for Development as a Point-of-Care Test. JAMA Pediatr. 2021 Jan 4:e205227. doi: 10.1001/jamapediatrics.2020.5227. Epub ahead of print. PMID: 33393977; PMCID: PMC7783591.

Pertea M, Kim D, Pertea GM, Leek JT, Salzberg SL. Transcript-level expression analysis of RNA-seq experiments with HISAT, StringTie and Ballgown. Nat Protoc. 2016 Sep;11(9):1650–67. doi: 10.1038/nprot.2016.095. Epub 2016 Aug 11. PMID: 27560171; PMCID: PMC5032908.

Pidsley R, Zotenko E, Peters TJ, Lawrence MG, Risbridger GP, Molloy P, Van Djik S, Muhlhausler B, Stirzaker C, Clark SJ. Critical evaluation of the Illumina MethylationEPIC BeadChip microarray for whole-genome DNA methylation profiling. Genome Biol. 2016 Oct 7;17(1):208. doi: 10.1186/s13059-016-1066-1. PMID: 27717381; PMCID: PMC5055731.

Poland GA, Ovsyannikova IG, Kennedy RB. Personalized vaccinology: A review. Vaccine. 2018 Aug 28;36(36):5350–5357. doi: 10.1016/j.vaccine.2017.07.062. Epub 2017 Jul 31. PMID: 28774561; PMCID: PMC5792371.

Price ME, Cotton AM, Lam LL, Farré P, Emberly E, Brown CJ, Robinson WP, Kobor MS. Additional annotation enhances potential for biologically-relevant analysis of the Illumina Infinium HumanMethylation450 BeadChip array. Epigenetics Chromatin. 2013 Mar 3;6(1):4. doi: 10.1186/1756-8935-6-4. PMID: 23452981; PMCID: PMC3740789.

Qiu S, He P, Fang X, Tong H, Lv J, Liu J, Zhang L, Zhai X, Wang L, Hu Z, Yu Y. Significant transcriptome and cytokine changes in hepatitis B vaccine non-responders revealed by genome-wide comparative analysis. Hum Vaccin Immunother. 2018 Jul 3;14(7):1763–1772. doi: 10.1080/21645515.2018.1450122. Epub 2018 May 14. PMID: 29580160; PMCID: PMC6067885.

Raeven RHM, van Riet E, Meiring HD, Metz B, Kersten GFA. Systems vaccinology and big data in the vaccine development chain. Immunology. 2019 Jan;156(1):33–46. doi: 10.1111/imm.13012. Epub 2018 Nov 13. PMID: 30317555; PMCID: PMC6283655.

Rohart F, Gautier B, Singh A, Lê Cao KA. mixOmics: An R package for ‘omics feature selection and multiple data integration. PLoS Comput Biol. 2017 Nov 3;13(11):e1005752. doi: 10.1371/journal.pcbi.1005752. PMID: 29099853; PMCID: PMC5687754.

Schillie SF, Murphy TV. Seroprotection after recombinant hepatitis B vaccination among newborn infants: a review. Vaccine. 2013 May 17;31(21):2506–16. doi: 10.1016/j.vaccine.2012.12.012. Epub 2012 Dec 17. PMID: 23257713.

Sendo S, Saegusa J, Morinobu A. Myeloid-derived suppressor cells in non-neoplastic inflamed organs. Inflamm Regen. 2018 Sep 17;38:19. doi: 10.1186/s41232-018-0076-7. PMID: 30237829; PMCID: PMC6139938.

Singh A, Shannon CP, Gautier B, Rohart F, Vacher M, Tebbutt SJ, Lê Cao KA. DIABLO: an integrative approach for identifying key molecular drivers from multi-omics assays. Bioinformatics. 2019 Sep 1;35(17):3055–3062. doi: 10.1093/bioinformatics/bty1054. PMID: 30657866; PMCID: PMC6735831.

Shannon CP, Blimkie TM, Ben-Othman R, Gladish N, Amenyogbe N, Drissler S, Edgar RD, Chan Q, Krajden M, Foster LJ, Kobor MS, Mohn WW, Brinkman RR, Le Cao KA, Scheuermann RH, Tebbutt SJ, Hancock REW, Koff WC, Kollmann TR, Sadarangani M, Lee AH. Multi-Omic Data Integration Allows Baseline Immune Signatures to Predict Hepatitis B Vaccine Response in a Small Cohort. Front Immunol. 2020 Nov 30;11:578801. doi: 10.3389/fimmu.2020.578801. PMID: 33329547; PMCID: PMC7734088.

Sharma M, Krammer F, García-Sastre A, Tripathi S. Moving from Empirical to Rational Vaccine Design in the ‘Omics’ Era. Vaccines (Basel). 2019 Aug 14;7(3):89. doi: 10.3390/vaccines7030089. PMID: 31416125; PMCID: PMC6789792.

Tenenhaus A, Philippe C, Guillemot V, Le Cao KA, Grill J, Frouin V. Variable selection for generalized canonical correlation analysis. Biostatistics. 2014 Jul;15(3):569–83. doi: 10.1093/biostatistics/kxu001. Epub 2014 Feb 17. PMID: 24550197.

Tripp CH, Ebner S, Ratzinger G, Romani N, Stoitzner P. Conditioning of the injection site with CpG enhances the migration of adoptively transferred dendritic cells and endogenous CD8+ T-cell responses. J Immunother. 2010 Feb-Mar;33(2):115–25. doi: 10.1097/CJI.0b013e3181b8ef5f. PMID: 20145551.

Tsang JS, Schwartzberg PL, Kotliarov Y, Biancotto A, Xie Z, Germain RN, Wang E, Olnes MJ, Narayanan M, Golding H, Moir S, Dickler HB, Perl S, Cheung F; Baylor HIPC Center; CHI Consortium. Global analyses of human immune variation reveal baseline predictors of postvaccination responses. Cell. 2014 Apr 10;157(2):499–513. doi: 10.1016/j.cell.2014.03.031. Erratum in: Cell. 2014 Jul 3;158(1):226. PMID: 24725414; PMCID: PMC4139290.

Tsang JS. Utilizing population variation, vaccination, and systems biology to study human immunology. Trends Immunol. 2015 Aug;36(8):479–93. doi: 10.1016/j.it.2015.06.005. Epub 2015 Jul 14. PMID: 26187853; PMCID: PMC4979540.

Tsang JS, Dobaño C, VanDamme P, Moncunill G, Marchant A, Othman RB, Sadarangani M, Koff WC, Kollmann TR. Improving Vaccine-Induced Immunity: Can Baseline Predict Outcome? Trends Immunol. 2020 Jun;41(6):457–465. doi: 10.1016/j.it.2020.04.001. Epub 2020 Apr 8. PMID: 32340868; PMCID: PMC7142696.

Villani AC, Satija R, Reynolds G, Sarkizova S, Shekhar K, Fletcher J, Griesbeck M, Butler A, Zheng S, Lazo S, Jardine L, Dixon D, Stephenson E, Nilsson E, Grundberg I, McDonald D, Filby A, Li W, De Jager PL, Rozenblatt-Rosen O, Lane AA, Haniffa M, Regev A, Hacohen N. Single-cell RNA-seq reveals new types of human blood dendritic cells, monocytes, and progenitors. Science. 2017 Apr 21;356(6335):eaah4573. doi: 10.1126/science.aah4573. PMID: 28428369; PMCID: PMC5775029.

Warimwe GM, Fletcher HA, Olotu A, Agnandji ST, Hill AV, Marsh K, Bejon P. Peripheral blood monocyte-to-lymphocyte ratio at study enrollment predicts efficacy of the RTS,S malaria vaccine: analysis of pooled phase II clinical trial data. BMC Med. 2013 Aug 21;11:184. doi: 10.1186/1741-7015-11-184. PMID: 23962071; PMCID: PMC3765422.

Wimmers F, Pulendran B. Emerging technologies for systems vaccinology - multi-omics integration and single-cell (epi)genomic profiling. Curr Opin Immunol. 2020 Aug;65:57–64. doi: 10.1016/j.coi.2020.05.001. Epub 2020 Jun 3. PMID: 32504952; PMCID: PMC7710534.

Wolf FA, Angerer P, Theis FJ. SCANPY: large-scale single-cell gene expression data analysis. Genome Biol. 2018 Feb 6;19(1):15. doi: 10.1186/s13059-017-1382-0. PMID: 29409532; PMCID: PMC5802054.

