## Supplemental figures and tables for "Machine learning-based single cell and integrative analysis reveals that baseline mDC predisposition predicts protective Hepatitis B vaccine response"

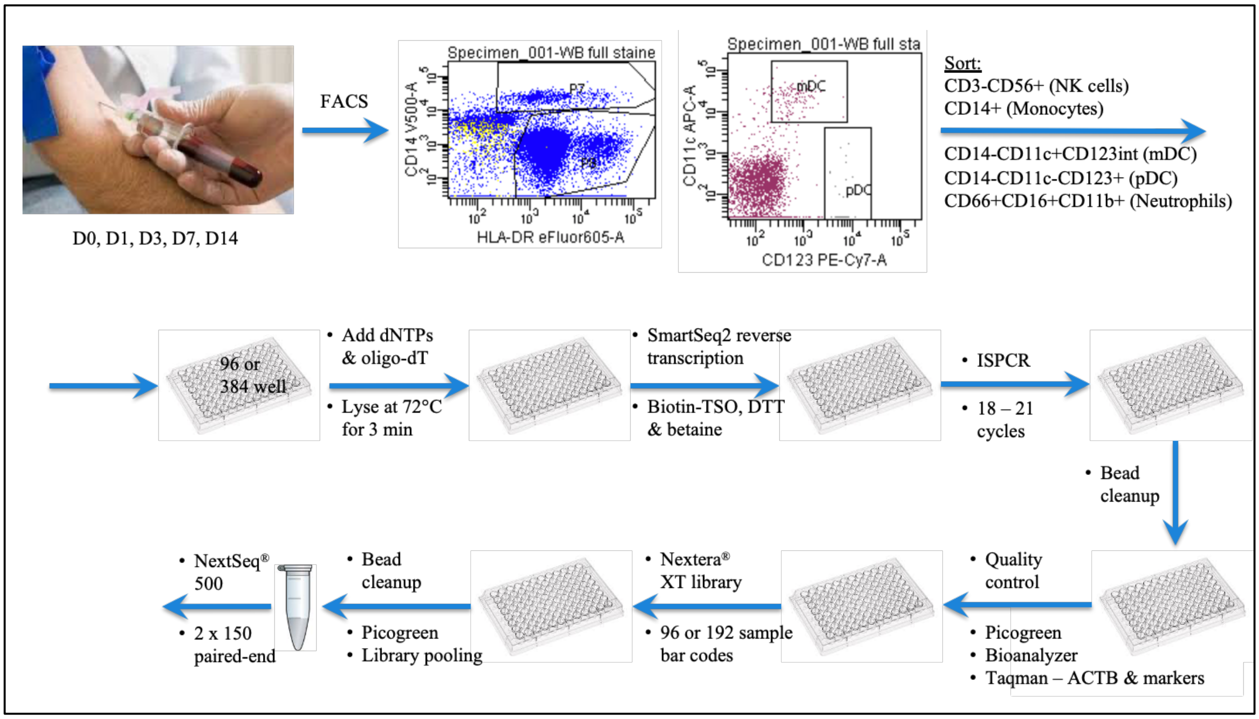


**Supplemental Figure 1. scRNAseq workflow -** A small cohort of human participants were challenges with the licensed Hepatitis B vaccine (*Engerix-B)*. Blood samples were collected before (Day 0) and Day 1, 3, 7 and 14 post-vaccination for single cell transcriptomics analysis of four different sorted innate immune cell subsets (NK cells, monocytes, mDC, and pDC) from three vaccine responders and three non-responders after one dose. Single cell RNA sequencing (scRNAseq) using a modified SmartSeq2 protocol was used for transcriptomics analysis.

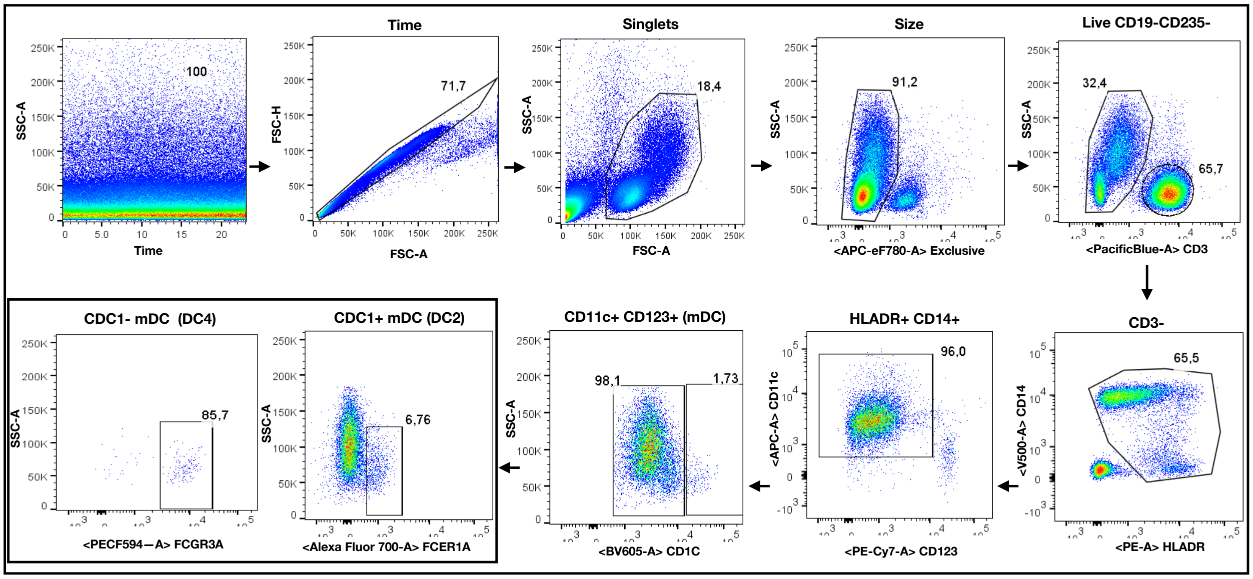


**Supplementary Figure 2. Gating strategy for DC2 and DC4 myeloid dendritic cells sorting -** The stability of the flow cytomery run was first checked by plotting the time versus scatter plot. Areas where there was poor flow were excluded and the "good flow gate" was used to identify the cell populations of interest in the panel. Stained PBMCs were first gated for singlets (FSC-H vs. FSCA) and then gated according to their size and granularity (SSC-A vs. FSC-A). The size gate was further analyzed for their uptakes of the live/dead fixable viability dye and to exclude cells expressing CD19 and CD325 surface markers. Live CD19-CD235- cells were further separated using CD3 surface marker to exclude T cells. The negative CD3 cells gate was carried over to identify CD14+ HLDR+ to further identify myeloid dendritic cells (mDC: CD11c+ CD123+/-). Finally, mDC cells were divided upon their CD1C expression to identify and sort two main mDC populations; DC2 cells (CDC1+ FCER1A) and DC4 (CDC1-FCGR3A+).

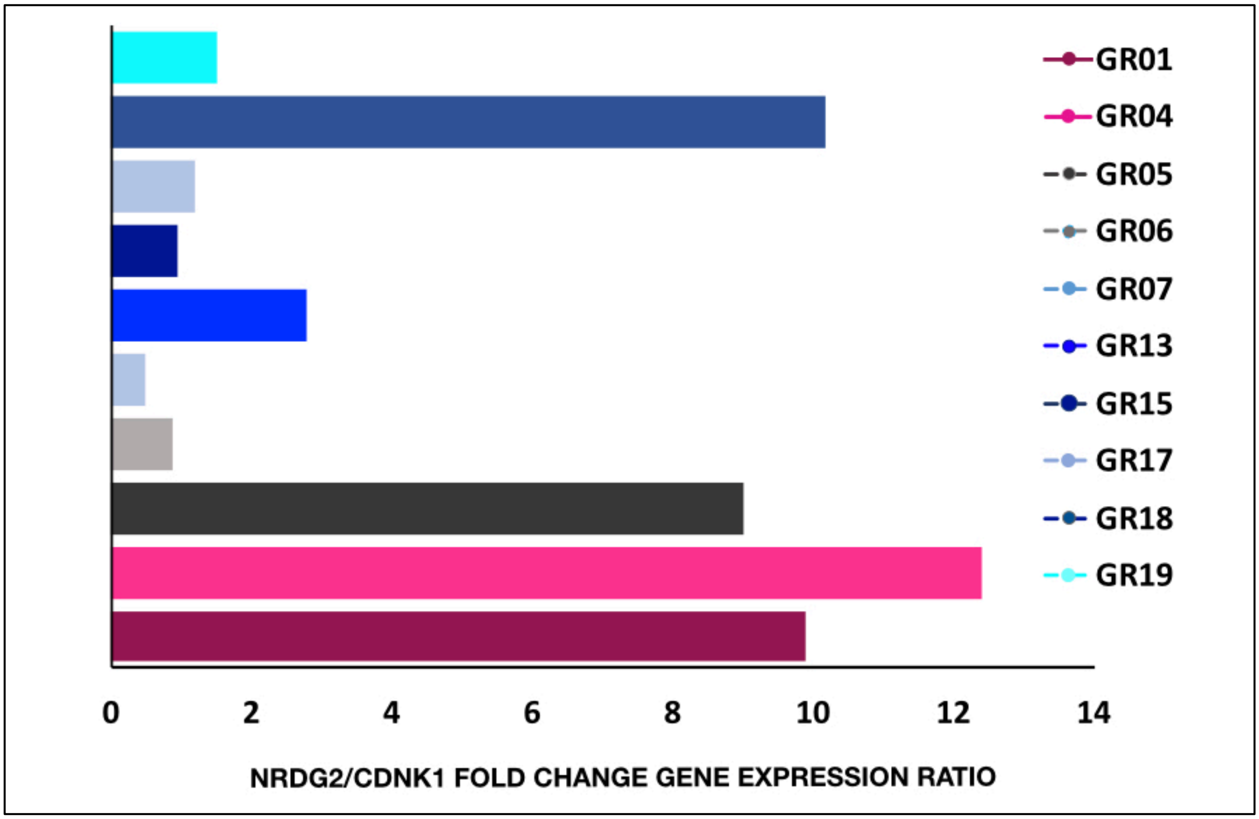


**Supplementary Figure 3. NDRG2 and CDNK1 gene expression in response to PolyIC at baseline differs between HBV vaccine early reponders and non-responders -** Whole blood collected from study participants prior to vaccination were incubated in Truculture tubes with or without PolyIC for 22hr at 37°C. At the end of the incubation time, plasma depleted blood was mixed to 1ml of trizol and total RNA extracted. Quantitative PCR was performed targeting NDRG2 and CDNK1 for all study participants shown in the figure. Data were normalised to the housekeeping gene (ACTB) and then to the unstimulated condition for each participant. Fold change of NDRG2 and CDNK1 normalized to ACTB expression (Delta Delta CT) for each participant were determined and the data are represented as ratios of the fold changes. Each coloured line graph shows the result of one study participant. The Y axis values were log transformed.

**Supplementary Table 1. DC2 and DC4 myeloid dendritic cells sorting panel** - Organization of panels for myeloid dendritic cells sorting from PBMC collected from peripheral whole blood. Nine-color panels were established in order to specifically sort mDC2 and mDC4 subsets as described in Supplementary Figure 2.

| Target | Fluor | Clone | Supplier | Cat. No. |
|---|---|---|---|---|
| Viability dye, CD19, CD235 | APC-eflour 780 | REA175, HIB19 | Mylteni, BD | 130-100-268 |
| FCER1A | Alex 700 | CRA-1 | Biolegend | 334630 |
| CD11c | APC | BU15 | eBio | 17-0138042 |
| FCGR3A ( CD16a) | PE-CF594 | 1001049 | R&D | FAB4325T |
| CD1C | BV605 | F10/21A3 | BD | 742748 |
| CD14 | BV480 | M5E2 | BD | 748304 |
| CD123 | PE-Cy7 | 6N6 | eBio | 25-1239 |
| HLADR | PE | LN3 | eBio | 12-995642 |
| CD3 | Pacific Blue | UCTH1 | BD | 558117 |

**Supplementary Table 2. CD4 and CD8 T cells proliferation staining panel** - Organization of panels for assessing autologous T cell proliferation in presence of either mDC4 or mDC2 dendritic cells. Cell proliferation was assessed by Oregon Green. The eight-color panel was established in order to quantify CD4 and CD8 proliferation after 5 days.

| Laser | Detector | Filter | Dichroic | Target | Fluorochrome | Clone | Supplier | Cat. No. |
|---|---|---|---|---|---|---|---|---|
| **Red (640 nm)** | A | 780/60 | 750LP | Viability dye | APC-eflour 780 | | eBio | 65-0865 |
| | B | 720/40 | 685LP | FCER1A | Alex 700 | CRA-1 | Biolegend | 334630 |
| **Green (532 nm)** | A | 780/60 | 750LP | CD8 | PE-Cy7 | RPA-T8 | eBio | 25-0087 |
| | D | 610/20 | 600LP | FCGR3A ( CD16a) | PE-CF594 | 1001049 | R&D | FAB4325T |
| | E | 585/15 | Blank | Ki67 | PE | B56 | BD | 556027 |
| | B | 525/50 | 505LP | OG | FITC | | Molecular Prob | 34550 |
| **Violet** | D | 610/20 | 595LP | CD4 | BV605 | RPA-T4 | BD | 562659 |
| | H | 450/50 | Blank | CD3 | Pacific Blue | OK3 | BD | 558117 |

**Supplemental Table 3. NS-Forest v2.0 marker genes for the seven Louvain clusters** - The celltype column gives the annotated name as described in the text. The f-measure column gives the discriminatory power of the markers as a set. The True Negative, False Positive, False Negative, and True Positive gives the confusion matrix results for single cell classification using the set of markers. The markerCount column gives the total number of markers in the set followed by the symbol identities of the gene expression markers.

| clusterName | cellType | f-measure | True Negative | False Positive | False Negative | True Positive | markerCount | 1 | 2 |
|---|---|---|---|---|---|---|---|---|---|
| Cluster_0 | pDC_DC6 | 0.98 | 1023 | 9 | 4 | 407 | 2 | JCHAIN | UGCG |
| Cluster_1 | NK | 1.00 | 1106 | 0 | 6 | 331 | 2 | PRF1 | CTSW |
| Cluster_2 | mDC_DC4 | 0.95 | 1173 | 0 | 56 | 214 | 2 | CDKN1C | LINC01272 |
| Cluster_3 | Monocytes | 0.89 | 1164 | 9 | 79 | 191 | 2 | CD14 | VCAN |
| Cluster_4 | mDC_DC2 | 0.85 | 1337 | 2 | 45 | 59 | 2 | NDRG2 | FCER1A |
| Cluster_5 | B cells | 0.98 | 1415 | 0 | 3 | 25 | 1 | CD79A | |
| Cluster_6 | pDC_DC5 | 0.95 | 1420 | 0 | 5 | 18 | 1 | AXL | |

**Supplemental Table 4. qRT-PCR results** – 96 mDCs were sorted into wells of a microtiter plate, cDNA prepared and qPCR for ACTB, CDKN1C and NDRG2 performed as described in the Methods section. The number of cells positive by qPCR is shown in the "count" columns. The percent of ACTB+ cells that were also positive for indicated marker gene is shown in the "%" columns. The relative proportion of NDRG2-expressing cells/CDKN1C-expressing cells is shown in the last column. Data for participant GR01 is shown as an example.

| Subject | timepoint | ACTB count | CDKN1C count | NDRG2 count | % CDKN1C mDC | % NDRG2 mDC | NDRG2/ CDKN1C |
|---|---|---|---|---|---|---|---|
| GR01 | Day 0 | 80 | 8 | 38 | 0.10 | 0.48 | 4.75 |
| GR01 | Day 1 | 51 | 33 | 13 | 0.65 | 0.25 | 0.39 |
| GR01 | Day 3 | 74 | 34 | 10 | 0.46 | 0.14 | 0.29 |
| GR01 | Day 7 | 49 | 9 | 8 | 0.18 | 0.16 | 0.89 |
| GR01 | Day 14 | 79 | 26 | 20 | 0.33 | 0.25 | 0.77 |

**Supplemental Table 5. Relative proportion of NDRG2-expressing cells/CDKN1C-expressing cells** – Relative proportion of NDRG2-expressing cells/CDKN1C-expressing cells were calculated as described in Supplemental Table 4 for all samples and all participant. Numbers in grey were interpolated from flanking timepoints because no cells were detected for at least one of the two mDC subsets in these samples.

| | Day0 | Day1 | Day3 | Day7 | Day14 |
|---|---|---|---|---|---|
| GR01 | 4.75 | 0.39 | 0.29 | 0.89 | 0.77 |
| GR04 | 1.50 | 0.22 | 0.37 | 0.52 | 1.20 |
| GR05 | 0.54 | 0.02 | 3.00 | 0.27 | 0.50 |
| GR06 | 2.00 | 0.71 | 1.59 | 2.47 | 0.92 |
| GR07 | 0.07 | 0.10 | 0.09 | 0.64 | 1.65 |
| GR13 | 0.12 | 0.34 | 0.24 | 0.28 | 0.25 |
| GR15 | 0.50 | 0.26 | 0.36 | 0.45 | 0.08 |
| GR17 | 0.11 | 0.16 | 0.21 | 0.10 | 0.28 |
| GR18 | 0.09 | 0.08 | 1.50 | 0.33 | 0.34 |
| GR19 | 0.29 | 0.59 | 0.96 | 1.33 | 1.39 |
